# AYUSH medicine as add-on therapy for mild category COVID-19; an open label randomised, controlled clinical trial

**DOI:** 10.1101/2020.12.06.20245019

**Authors:** Anusha Rao, R Ranganatha, G Vikneswaran, C Sagar, R Mathu, M Sherin, Alben sigamani, M. Ravi Kumar Reddy

## Abstract

**Background:** AYUSH formulations have a potential role in symptomatic treatment, preventing disease progression and improving quality of life in COVID-19 patients.

**Objective:** To study the effect of AYUSH formulation (Kabasura Kudineer tablets, Shakti drops and Turmeric plus) as an add-on treatment in patients with mild COVID - 19

**Methodology:** Single centre, two arms, open labelled randomized controlled trial with a total of 30 patients (15 in the intervention arm and 15 in the standard care arm). Intervention arm received a combination of 3 AYUSH formulation along with the standard of care treatment for 21 days. All patients were followed for 28 days. Symptom severity (using Modified Jackson scale), negative conversion of SARS-CoV-2 RNA (using RTPCR) and quality of life (WHOWOL BREF questionnaire) was assessed.

**Results:** Fifteen patients (93.8%) in the intervention group and twelve patients (92.3%) in the standard care arm had complete resolution of symptoms (P value= 0.36). Negative conversion for SARS-CoV-2 was seen in thirteen patients (92.9%) in intervention arm and eleven patients (100%) in standard care arm at day 28 (P value = 0.56). There was no difference in the quality of life scores between the 2 groups.

**Conclusion:** The use of Ayush interventions as add-on therapy did not negatively impact the clinical outcomes in COVID-19. This trial confirmed the safety and tolerability of Kabasura Kudineer tablets, Shakti drops and Turmeric plus tablets when used use among mild to moderate symptom category, of COVID-19. There were no serious adverse events in the treated group. There was no clinical progression of disease from baseline status and all trial participants recovered fully by day 28. A longer follow up and a larger sample size is recommended for future definitive trials with this alternative medicine (AYUSH) combination.

## Introduction

Novel corona virus disease (COVID-19) has become a pandemic, causing global health crisis. The condition is caused by severe acute respiratory syndrome corona virus 2 (SARS-CoV-2) transmitted from humans to humans through aerosols. (1) Clinical spectrum of COVID-19 manifestation ranges from asymptomatic presentation to pneumonia, ARDS, multiorgan dysfunction and death. (2,3) Based on the severity, the disease is classified as mild, moderate, severe and critical. (4) Mild form accounts for approximately 80% of the COVID illness. Most common symptoms, reported by a WHO-China Joint Commission report based on over fifty thousand cases were, fever (87.9%), dry cough (67.7%), fatigue (38.1%), sputum production (33.4%), and shortness of breath (18.6%). (5)

The current treatment strategy for COVID-19 is largely symptomatic depending on the severity of illness such as paracetamol, antitussives, nutritional supplement, oxygen therapy, glucocorticoids and ECMO. (6) Ayurveda has unique methods of approaching a newly detected disease. Rather than focusing on the microbiological etiology, Ayurveda embraces a holistic technique for elaborating the details of the disease at hand, with a three-pointed approach in the elaboration of an unknown disease-the natural history of the disease (Vikara Prakriti), the site of the pathological process (Adhishtanam/Srotasa) and etiological features (Samuthaana Visesham). The Ministry of AYUSH has also recently published guidelines regarding use of different formulations for symptom relief in COVID-19. (7)

Evidences show that herbal medicines have been successfully used in the treatment of SARS COV infection. (8) A Cochrane review has shown that herbal medicines when combined with western medicines may help in improvement of symptoms and quality of life in patients with SARS. (9) Thus, the present study was undertaken to study the effect of a combination of 3 AYUSH medicines (Kabasura Kudineer tablets, Shakti drops and Turmeric plus tablets) on symptom improvement, disease progression and quality of life in patients with mild COVID-19

## Methodology

The study was initiated after ethics committee approval (NHMEC S38/2020) and informed consent was obtained from all study participants. All study related activities were conducted in accordance with the AYUSH (GCP-ASU) Guidelines for Good Clinical Practice, the ICMR National Ethical Guidelines for Bio medical and Health research involving human participants. This trial is being reported as per the CONSORT checklist. The study was registered in the Clinical trial registry of India (CTRI/2020/07/026371)

### Eligibility criteria

For the study we included uncomplicated cases of COVID-19 infection of either gender between 20 to 55 years of age, with no associated co-morbid illness. We excluded patients on immunosuppressive therapy, pregnant and lactating mothers and those with advanced stage of illness requiring emergency medical interventions.

### Study design and intervention details

The study was a single centre, open label. randomized controlled trial including a total of 30 patients with mild COVID symptoms. Patients were classified as Stage 1 and stage 2 depending on the lung symptoms. Stage 1 included patients with symptoms of headache, sore throat, fever, slight cough, malaise, and fatigue. Stage 2 included patients with cough, breathlessness, and pain in the chest with fever in addition to the symptoms mentioned in stage 1. Eligible patients were screened from the COVID ward. After obtaining informed consent patients were randomized into either standard care only or standard care with AYUSH medicines.

Study data were collected and managed using REDCap electronic data capture tools hosted at [Narayana Hrudayalaya Limited Bangalore]. (10,11) Randomization was done by Interactive Web based Randomisation Service (IWRS) on REDCap.

Patients in the standard care received paracetamol, antitussives, vitamin C, Zinc, antibiotics and ivermectin. In addition, patients in the intervention arm received Kabasura Kudineer tablets (2 tablets taken thrice a day before food) and Shakti drops (6 drops with 100 ml of water thrice a day before food) for stage 1; Kabasura Kudineer tablets (2 tablets taken thrice a day before food) and Shakti drops (6 drops with 100 ml of water thrice a day before food) and Turmeric plus tablets (2 tablets thrice a day after food) for stage 2. All the 3 formulations are GMP approved. Along with this, patients were advised dietary and lifestyle modifications such as consuming freshly prepared easily digestible, light diet; avoid using sweets and food which is heavy to digest, chilled, fried, oily, fermented food items, cold and refrigerated food /beverages.

### Outcome Assessment

Standard care was provided until symptom resolved and patient condition improved. AYUSH medicines were given for a period of 21 days. Patients were monitored daily for improvement of symptoms, development of any adverse effects or disease progression until discharge followed by telephonic assessment at day 28. Symptom severity was assessed using Modified Jackson symptom severity score. Eight symptoms namely sneezing, nasal discharge, nasal congestion, sore throat, cough, headache, malaise, and fever were scored from 0 to 3 (0 for no symptoms, 1 for mild, 2 moderate and 3 for severe symptoms). (12) Patients were asked to rate their symptoms based on the severity. Quality of life was assessed using WHO QOL BREF questionnaire. It is a self-administered questionnaire with 26 items under 4 main domains (physical health, psychological health, social relationship and environment). Each item was given a score of 1 to 5 (1 when “very dissatisfied” and 5 when “very satisfied”). (13) RT PCR to test for SARS-Cov-2 negative conversion was done at day 14 and repeated weekly till day 28 till they turn negative.

Safety assessment was done based on symptom assessment, clinical examination and blood investigations. Blood investigations included complete blood count, blood sugar level, liver and renal function test performed at baseline, day 14 and day 21.

### Sample size and statistical analysis

A total of 30 patients were planned for the study (15 in the standard care only and 15 in AYUSH plus standard care). Descriptive statistics was used for baseline characteristics. Continuous variables were expressed using mean and standard deviation. Categorical variables were expressed using frequency and percentage. Proportion of patients achieving improvement in symptoms at day-7 was compared between 2 groups using Fischer exact test. Improvement in quality of life between the 2 groups on day 28 was compared using Mann Whitney U test. P value less than 0.05 was considered statistically significant.

## Results

A total of 30 patients were enrolled for the study (15 in each group). One patient was wrongly randomized into AYUSH plus standard arm making it 16 in the intervention arm and 14 in the standard care arm. (Figure-1). Baseline characteristics were shown as per randomization and all patient characteristics were comparable between the 2 groups. The mean age was 27.2 ± 6.81 in the intervention arm and 28.20 ± 4.96 in the standard care arm. The predominant symptoms were fever (26.7% vs 33.3%), cough (26.7% vs 40%), sore throat (53.3% vs 40%), running nose (26.7% vs 33.3%), general weakness (66.7% vs 40%), and headache (33.3% vs 33.3%) in the intervention group and standard care group respectively. (Table-1)

**Figure 1.**
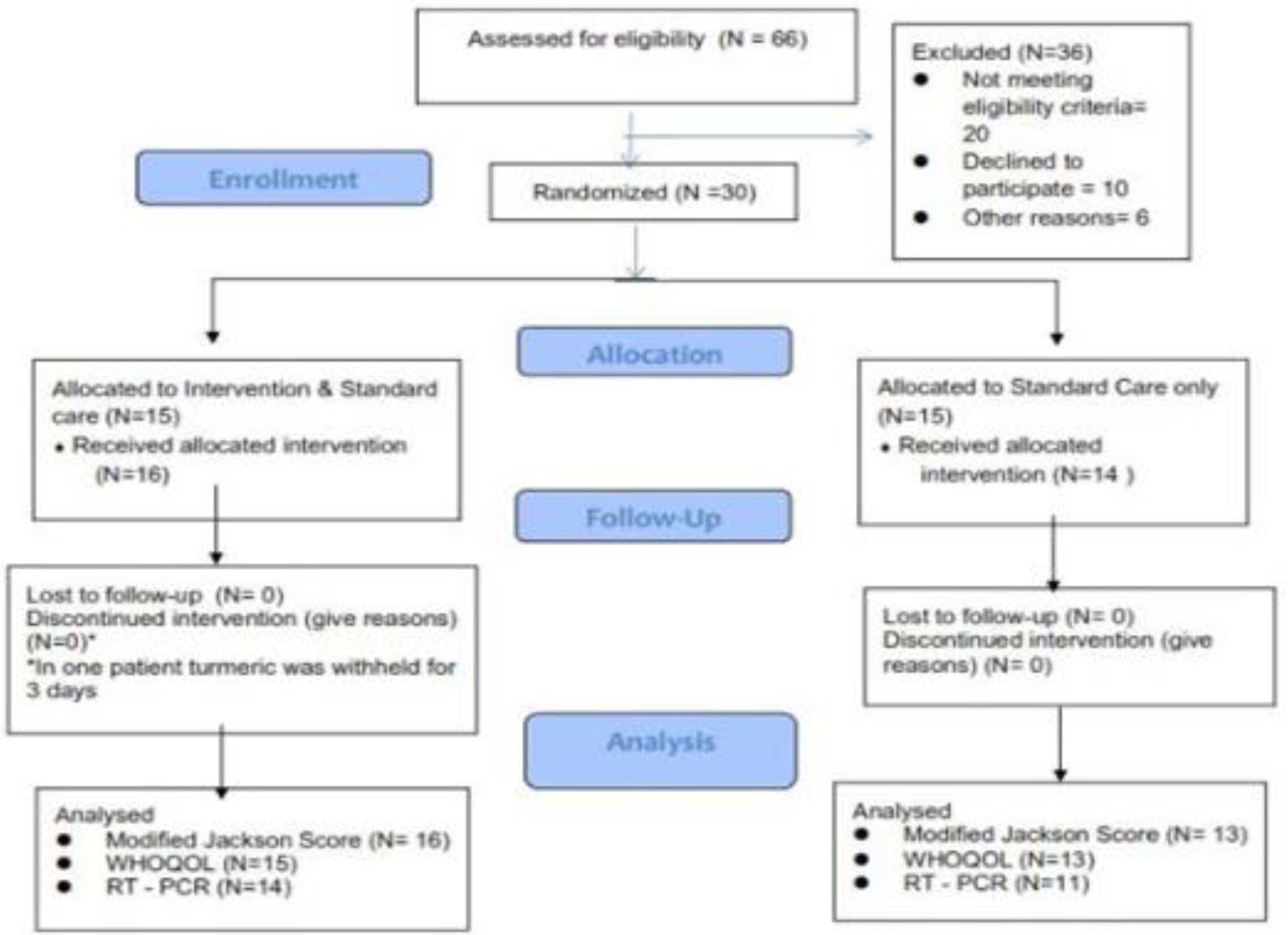
Study flow diagram.

**Table-1.**
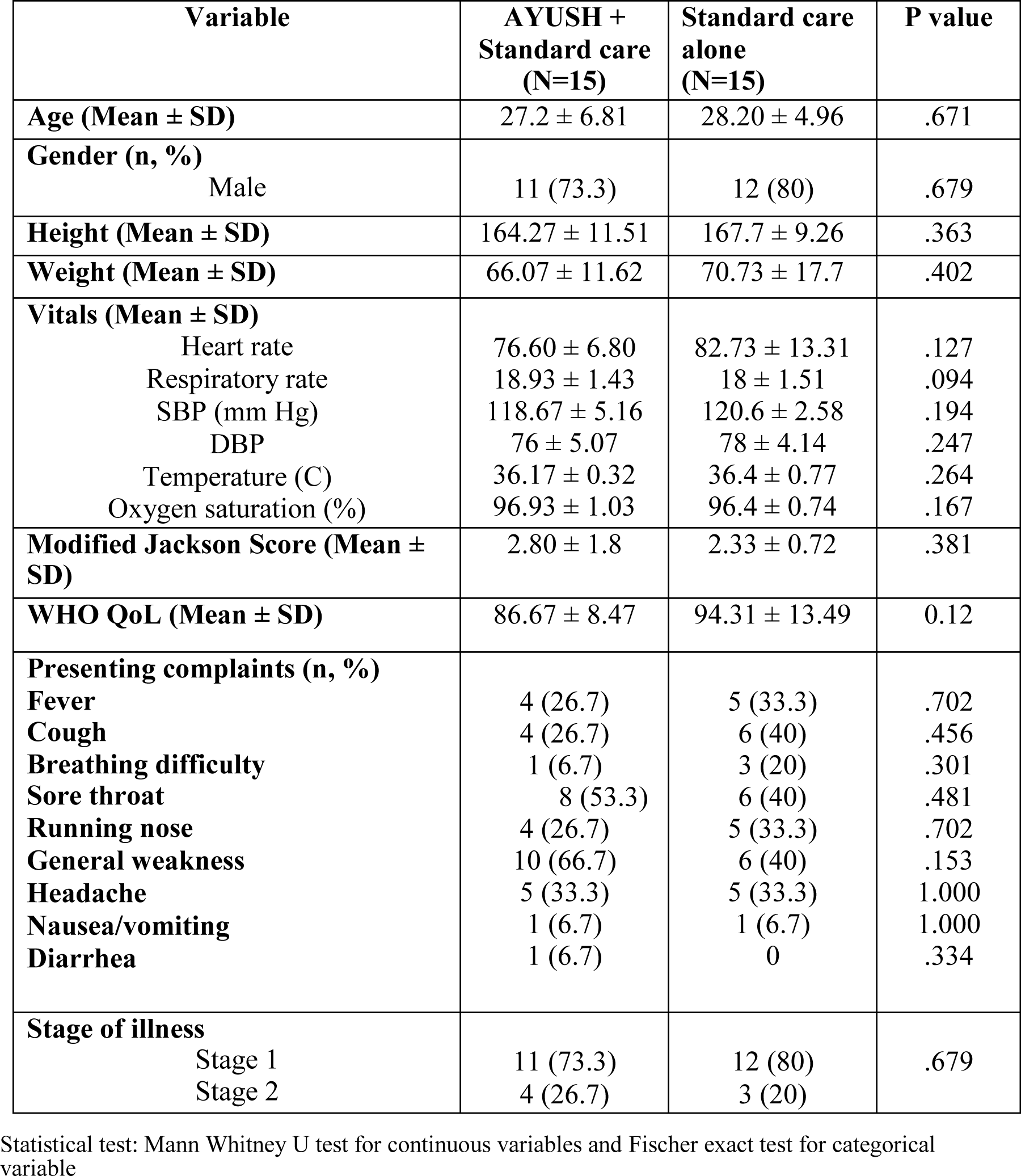
Baseline characteristics of study population.

| **Variable** | **AYUSH +**<br>**Standard care**<br>**(N=15)** | **Standard care**<br>**alone**<br>**(N=15)** | **P value** |
| --- | --- | --- | --- |
| **Age (Mean ± SD)** | 27.2 ± 6.81 | 28.20 ± 4.96 | .671 |
| **Gender (n, %)**<br>Male | 11 (73.3) | 12 (80) | .679 |
| **Height (Mean ± SD)** | 164.27 ± 11.51 | 167.7 ± 9.26 | .363 |
| **Weight (Mean ± SD)** | 66.07 ± 11.62 | 70.73 ± 17.7 | .402 |
| **Vitals (Mean ± SD)** |  |  |  |
| Heart rate | 76.60 ± 6.80 | 82.73 ± 13.31 | .127 |
| Respiratory rate | 18.93 ± 1.43 | 18 ± 1.51 | .094 |
| SBP (mm Hg) | 118.67 ± 5.16 | 120.6 ± 2.58 | .194 |
| DBP | 76 ± 5.07 | 78 ± 4.14 | .247 |
| Temperature (C) | 36.17 ± 0.32 | 36.4 ± 0.77 | .264 |
| Oxygen saturation (%) | 96.93 ± 1.03 | 96.4 ± 0.74 | .167 |
| **Modified Jackson Score (Mean ± SD)** | 2.80 ± 1.8 | 2.33 ± 0.72 | .381 |
| **WHO QoL (Mean ± SD)** | 86.67 ± 8.47 | 94.31 ± 13.49 | 0.12 |
| **Presenting complaints (n, %)** |  |  |  |
| **Fever** | 4 (26.7) | 5 (33.3) | .702 |
| **Cough** | 4 (26.7) | 6 (40) | .456 |
| **Breathing difficulty** | 1 (6.7) | 3 (20) | .301 |
| **Sore throat** | 8 (53.3) | 6 (40) | .481 |
| **Running nose** | 4 (26.7) | 5 (33.3) | .702 |
| **General weakness** | 10 (66.7) | 6 (40) | .153 |
| **Headache** | 5 (33.3) | 5 (33.3) | 1.000 |
| **Nausea/vomiting** | 1 (6.7) | 1 (6.7) | 1.000 |
| **Diarrhea** | 1 (6.7) | 0 | .334 |
| **Stage of illness** |  |  |  |
| Stage 1 | 11 (73.3) | 12 (80) | .679 |
| Stage 2 | 4 (26.7) | 3 (20) |  |

Statistical test: Mann Whitney U test for continuous variables and Fischer exact test for categorical variable

Data were analysed using modified intention to treat analysis (patients received at least one dose of the study drug and available for analysis). All patients completed the trial and discharged in stable condition. There was no loss to follow up by the end of 28 days. Modified Jackson score was analysed in 29 patients at day 7 (one not willing to continue) and WHOQOL score analysed in 28 patients (2 patients not willing for assessment). Fifteen patients in the intervention group and 12 patients in the standard group had complete resolution of symptoms by day 7. There was no significant difference in terms of symptom improvement between the 2 groups. Fifteen patients (93.8%) in the intervention group and twelve patients (92.3%) in the standard care arm had complete resolution of symptoms (P value= 0.36) (Table-3). Though both the groups showed improvement in quality-of-life score, there was no significant difference in improvement of quality of life between the 2 groups. (Table-4)

**Table-2.** Baseline blood parameters.

| **Blood Investigations** | **AYUSH + Standard care**<br>**(N=15)** | **Standard care**<br>**(N=15)** | **P**<br>**value** |
| --- | --- | --- | --- |
| **Haematology** |  |  |  |
| Haemoglobin (gm/dl) | 14.1 ± 3.04 | 15.3 ± 1.38 | 0.17 |
| TLC (per mcl) | 6.56 ± 2.33 | 5.9 ± 1.8 | 0.44 |
| Neutrophils(%) | 54.3 ± 6.38 | 51.6 ± 12.7 | 0.51 |
| Lymphocytes(%) | 34.4 ± 8.5 | 34.6 ± 11.5 | 0.95 |
| Monocytes(%) | 9.13 ± 1.96 | 10.7 ± 4.5 | 0.25 |
| Eosonophils(%) | 1.6 ± 1.5 | 2.5 ± 2.9 | 0.32 |
| Basophils (%) | 0.44 ± 0.29 | 0.42 ± 0.19 | 0.80 |
| **Platelet (n, %)** |  |  |  |
| 1,00,000 to 1,50,000 | 1 (6.7) | 0 | 1.00 |
| >1,50,000 | 13 (86.7) | 14 (93.3) |  |
| Not available | 1 (6.7) | 1 (6.7) |  |
| **Biochemical tests** |  |  |  |
| RBS (mg/dl) | 95.3 ± 30.2 | 94.9 ± 25.0 | 0.97 |
| Blood Urea (mmol/l) | 8.6 ± 2.5 | 9.7 ± 2.4 | 0.28 |
| Serum Creatinine<br>(mg/dl) | 0.74 ± 0.19 | 0.72 ± 0.17 | 0.70 |
| **SGOT (n, %)** |  |  |  |
| Normal | 12 (80) | 11 (73.3) | 1.00 |
| 1 to 2 times upper<br>normal limit | 1 (6.7) | 2 (13.3) |  |
| Not available | 2 (13.3) | 2 (13.3) |  |
| **SGPT (n, %)** |  |  |  |
| Normal | 8 (53.3) | 8 (53.3) | 1.00 |
| 1 to 2 times upper<br>normal limit | 5 (33.3) | 5 (33.3) |  |
| Not available | 2 (13.3) | 2 (13.3) |  |

Statistical test used: Fischer exact test for categorical variables and Mann Whitney U test for continuous variables.

**Table-3.** Comparison of improvement in symptom severity using Modified Jackson score (day 7)

|  | AYUSH plus<br>standard care<br>(N=16) | Standard care alone<br>(N=13) | P value |
| --- | --- | --- | --- |
| Patients achieved<br>resolution of<br>symptoms | 15 (93.8%) | 12 (92.3%) | 0.36 |

Statistical test used Fischer exact test

**Table-4.** Comparison of Quality-of-Life score.

|  |  | AYUSH plus<br>standard care<br>(N= 15) | Standard care<br>alone<br>(N=13) | P value |
| --- | --- | --- | --- | --- |
| QOL score<br>total | Baseline | 86.67 ± 8.47 | 94.31 ± 13.49 | 0.12 |
| | 28 days | 95.47 ± 9.62 | 106.38 ± 16.89 | 0.12 |
| | Difference | 8.8 ± 12.35 | 12.07 ± 18.08 | 1 |
| Physical<br>health | Baseline | 22.93 ± 2.91 | 25.69 ± 4.25 | 0.246 |
| | 28 days | 24.73 ± 2.21 | 29.38 ± 4.66 | 0.007 |
| | Difference | 1.8 ± 3.44 | 3.69 ± 5.03 | 1 |
| Psychologic<br>al health | Baseline | 20.33 ± 2.49 | 21.85 ± 3.76 | 0.44 |
| | 28 days | 22.73 ± 3.15 | 24.31 ± 4.64 | 1 |
| | Difference | 2.4 ± 3.83 | 2.46 ± 4.29 | 0.71 |
| Social<br>relationship | Baseline | 10.47 ± 1.59 | 11 ± 1.41 | 0.7 |
| | 28 days | 11.20 ± 2.17 | 12.08 ± 1.84 | 1 |
| | Difference | 0.7 ± 2.49 | 1.07 ± 2.01 | 1 |
| Environmen<br>t<br>factors | Baseline | 25.67 ± 2.92 | 28.08 ± 4.48 | 0.24 |
| | 28 days | 28.87 ± 3.66 | 32 ± 6.12 | 0.05 |
| | Difference | 3.2 ± 3.98 | 3.92 ± 5.85 | 1 |

Statistical test used: Mann Whitney U test

RTPCR testing at 14 days showed 6 patients (54.5%) in intervention arm and 7 patients (70%) in standard care arm negative for COVID (P value 0.39). By day 28, thirteen patients (92.9%) in intervention arm and 11 patients (100%) in standard care arm were negative (P value 0.56 Table-5).

**Table-5.**
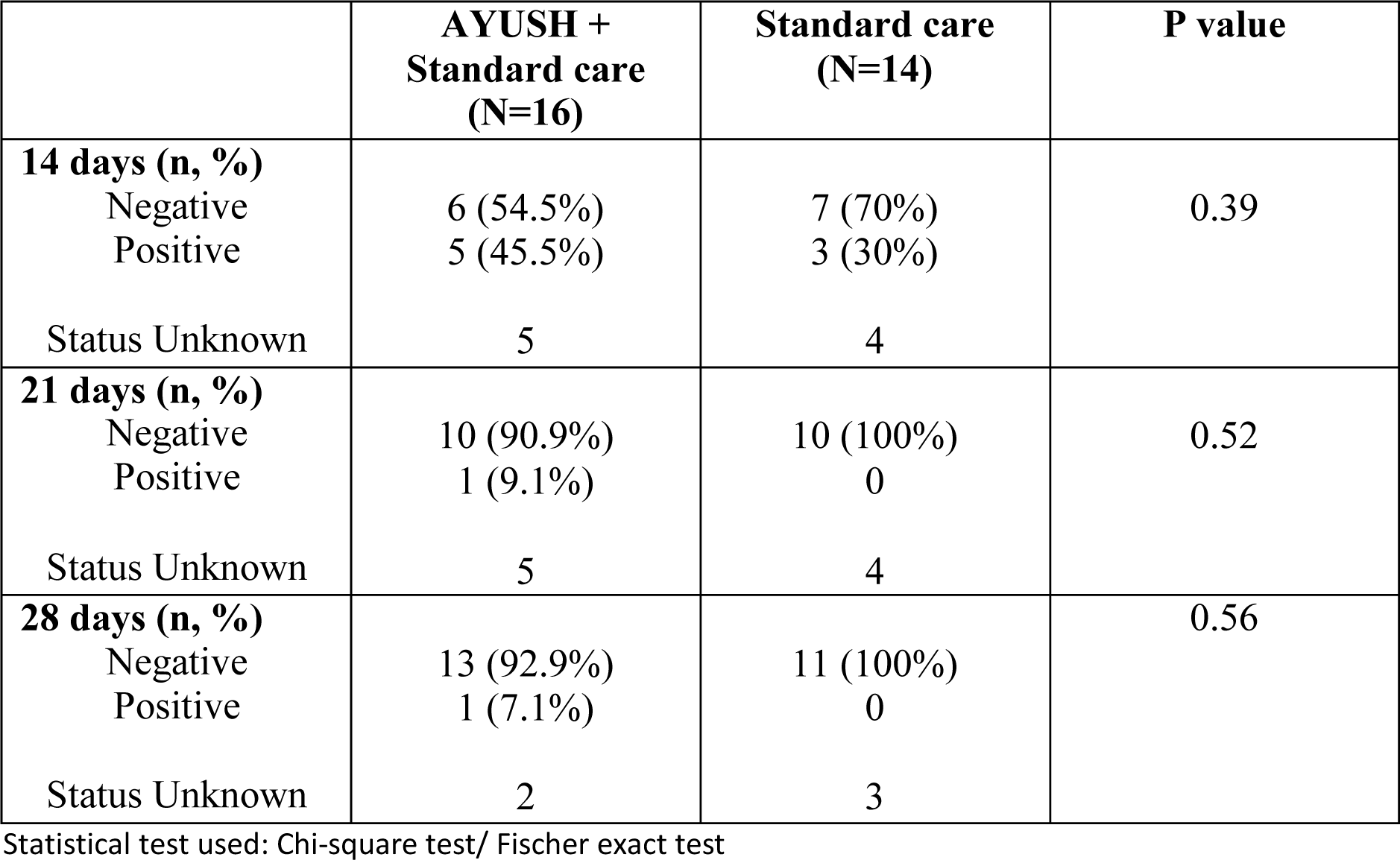
Comparison of SARS CoV-2 negative conversion between two groups.

Overall, the study participants had good compliance to study drugs. Turmeric plus tablet was withheld in one of the patients for 3 days as the participant was having menorrhagia. Excluding her the compliance for turmeric plus comes to 90.46% Compliance for Kabasura Kudineer tablets and Shakti drops were 86.76% and 91.08% respectively.

### Adverse events

One participant in the standard of care alone arm, required oxygen support on experiencing difficulty in breathing. This serious adverse event was recorded and reported to ethics committee. Three participants in the treatment group (AYUSH plus Standard of Care) experienced non related adverse events; one had uveitis, the other hyperglycaemia and one person experienced menorrhagia. They did not qualify to be reported as serious adverse events.

## Discussion

Study was conducted to evaluate the efficacy of a combination of AYUSH medicines along with standard of care versus standard of care alone on improvement in symptom severity, conversion to COVID-19 negative status and improvement in quality of life in patients with mild COVID infection. Patients with mild COVID illness were divided into stage 1 and stage 2 depending on the presence or absence of lung symptoms (such as cough, breathless, pain in the chest while breathing etc). Patients in stage 1 received Kabasura Kudineer tablets and Shakti drops along with the standard of care. Patients in stage 2 received Kabasura Kudineer tablets, Shakti drops and Turmeric plus tablets along with the standard of care.

Kabasura Kudineer tablet containing 15 active ingredients has potent antiviral and immunomodulatory properties and is recommended in the Siddha guidelines for the treatment of mild and moderate COVID symptoms (7,10) Shakti drops contain a combination of 8 Ayurvedic ingredients (Amla, Ashwagandha, Bhringaraj, Brahmi, Amruth, Shankapushpi, Shatavari and Yashtimadhu).(11) The combination supports immune function, strengthens respiration and rejuvenates the body. Turmeric plus is a combination of turmeric and black pepper. Curcumin, active component of turmeric is responsible for its anti-inflammatory effect, antiseptic and antibacterial properties and black pepper acts as a bioenhancer. (12, 13)

For analysis of symptom resolution, patients with a score of “zero” for all the symptoms in the Modified Jackson symptom severity score was considered. Both the groups had complete resolution of symptoms by day 14. There was no significant difference in terms of symptom resolution on day 7 (93.8% in the intervention vs 92.3% in the standard care. P value 0.36) For SARS COV-2 negative conversion, RT-PCR with sample collected using throat swab was used. The study did not show any significant difference in terms of negative conversion rate between the intervention arm and standard care both at day 14 (54.5% vs 70%, P value 0.39) as well as day 28 (92.9% vs 100%, P value 0.56) respectively.

Quality of life assessed using WHO QOL BREF questionnaire showed improvement in all the 4 domains (physical health, psychological health, social relationship and environmental factors) by day 28 in both the groups. However, intervention group did not significantly improve the quality-of-life score when compared with the standard care. Patients with COVID-19 show reduced quality of life score especially in the physical and psychological domain. The factors influencing quality of life are age, duration of hospital stay and presence of persistent symptoms such as myalgia post discharge. (14,15)

The patients in our study had low symptom severity score at baseline and showed complete resolution of symptoms by day 14. This may probably be the reason for the lack of improvement seen in the quality of life between the 2 groups. Also since this was a pilot study, the number of patients were small. These are the limitations of our study.

## Conclusion

This trial concludes that a combination Kabasura Kudineer tablets, Shakti drops and Turmeric plus tablets used as an add on along with standard of care treatment of COVID-19 is safe. All the participants who received the combination did not experience clinical worsening of their illness. It can be considered as a safe alternative medicine add on therapy for COVID-19. Further studies are needed to evaluate the long term benefits in COVID-19.

## Data Availability

All data referred to in the manuscript are available in REDCap electronic data capture tools
hosted at [Narayana Hrudayalaya Limited Bangalore].

